# Variation and impact of polygenic hematological traits in monogenic sickle cell disease

**DOI:** 10.1101/2022.01.26.22269309

**Authors:** Thomas Pincez, Ken Sin Lo, Anne-Laure Pham Hung d’Alexandry d’Orengiani, Melanie E. Garrett, Carlo Brugnara, Allison E. Ashley-Koch, Marilyn J. Telen, Frédéric Galactéros, Philippe Joly, Pablo Bartolucci, Guillaume Lettre

## Abstract

Several complications observed in sickle cell disease (SCD) are influenced by variation in hematological traits (HT), such as fetal hemoglobin (HbF) level and neutrophil count. Previous large-scale genome-wide association studies carried out in largely healthy individuals have identified 1000s of variants associated with HT, which have then been used to develop multi-ancestry polygenic trait scores (PTS). Here, we tested if these PTS associate with HT in SCD patients and can improve the prediction of SCD-related complications. In 2,056 SCD patients, we found that the PTS predicted less HT variance than in non-SCD African-ancestry individuals. This was particularly striking at the Duffy/*DARC* locus, where we observed an epistatic interaction between the SCD genotype and the Duffy null variant (rs2814778) that led to a two-fold weaker effect on neutrophil count. PTS for these routinely measured HT were not associated with complications in SCD. In contrast, we found that a simple PTS for HbF that includes only six variants explained a large fraction of the phenotypic variation (17.1-26.4%), associated with acute chest syndrome and stroke risk, and improved the prediction of vaso-occlusive crises. Using Mendelian randomization, we found that increasing HbF by 4.8% reduces stroke risk by 36% (*P* = 0.0008). Taken together, our results highlight the importance of validating PTS in large diseased populations before proposing their implementation in the context of precision medicine initiatives.

## INTRODUCTION

Polygenic trait scores (PTS) have been developed in an effort to harness the power of large-scale human genetic studies to make useful clinical predictions, and many studies already support their value in the context of precision medicine initiatives.^1,2^ One abundantly discussed limitation of PTS is their poor performance when tested in populations that have different ancestral backgrounds than the populations in which they were optimized.^3^ Another equally important aspect that has not been as extensively studied is how well PTS, which are normally calibrated in “healthy” individuals, perform in “diseased” individuals.^4^ This is important because PTS could, in theory, be useful to stratify patients into mild or severe categories. For instance, a PTS for estimated glomerular filtration rate (eGFR) could help identify hypertensive patients more likely to suffer from kidney failure, but because this PTS_eGFR_ would have been developed in largely normotensive individuals, it is unclear how useful it would be in patients with hypertension.

To address this question, we took advantage of the classic monogenic disorder sickle cell disease (SCD), and explored how this pathology impacts the performance of PTS for hematological traits (HT). SCD, the most frequent monogenic disease worldwide, is caused by mutations in the β-globin gene.^5^ Despite being primarily a mature red blood cell disorder, SCD is associated with a broad range of consequences, both hematological (*e*.*g*. hemolysis, ineffective erythropoiesis) and extra-hematological (*e*.*g*. inflammatory state, endothelial cell activation).^5,6^ SCD patients present a wide range of complications such as vaso-occlusive crisis (VOC), acute chest syndrome (ACS), stroke, and end-organ dysfunction, and their life expectancy is reduced when compared to the general population.^5^ Critically, the causes of this clinical heterogeneity are only poorly understood.

HT are among the main factors known to be associated with clinical outcomes in SCD. fetal hemoglobin (HbF) is a major disease modifier, associated with reduction in the occurrence of several complications such as VOC, ACS and death.^7–9^ A percentage of HbF > 30% is associated with an almost complete absence of complications in SCD patients.^10,11^ However, most SCD patients have lower HbF levels while not receiving disease-modifying therapy and the risk reduction associated with HbF has not been quantified in large cohorts for some complications such as stroke. Several other HT have been associated with SCD-related complications, notably elevated white blood cell (WBC) and neutrophil counts with survival,^7,12,13^ low hemoglobin levels with composite severe outcomes and death,^13,14^ and platelet count with ACS.^15^

Here, we investigated if PTS for HT developed in largely healthy cohorts also associate with blood-cell phenotypes in SCD patients. Furthermore, we evaluated the clinical relevance of these PTS in terms of predicting complications in this patient population.

## SUBJECTS AND METHODS

### Populations

We collected data from three SCD cohorts with genome-wide genotype information available: the Cooperative Study of Sickle Cell Disease (CSSCD, n = 1278),^16^ Genetic Modifier (GEN-MOD, n = 406),^17–19^ and Mondor/Lyon (n = 372)^20^ (**Table S1**). Data collection was made according to the Helsinki declaration and the study was approved by the institutional ethics committees. DNA genotyping has been described elsewhere^18,21^ and we used reference haplotypes from the TOPMed project^22^ to impute missing genotypes. Eight to 14 hematological traits were measured at steady state and available in these SCD cohorts. All patients were at > 3 months from a blood transfusion and only ten patients were taking hydroxyurea at baseline. For comparison, we also accessed data from African-ancestry individuals from the BioMe cohort^23,24^ and the UK Biobank.^25^

### PTS for HT

For all HT except HbF, we used the multi-ancestry PTS derived by the Blood-Cell Consortium to test for association with HT.^23^ Briefly, these PTS considered the effect sizes of variants that reach genome-wide significance in multi-ancestry meta-analyses of 746,667 individuals, including 15,171 African-ancestry participants. We generated additive PTS for each individual and HT by calculating the sum of HT-increasing alleles weighted by the corresponding genome-wide association study (GWAS) effect size. For HbF, we derived a PTS by considering the conditional effect sizes of six variants at three loci (*BCL11A, HBS1L-MYB, HBB*) associated with HbF levels in SCD patients.^26–28^ We tested the association between normalized PTS and HT by linear regression with the four first principal components as covariables.

### GWAS of HT in SCD patients

We adjusted HT for sex and age, and then applied inverse normal transformation. We performed GWAS for each HT available in the three SCD cohorts separately using RvTests (v20190205),^29^ testing an additive genetic model and correcting for the 10 first principal components. We then performed a meta-analysis of the GWAS results using METAL.^30^

For the Duffy/*DARC* null variant (rs2814778) association with neutrophil and WBC counts, we compared the additive and recessive models in each cohort, correcting each model for the 10 first principal components. We calculated the effect of rs2814778 on raw neutrophil and WBC counts using a recessive model with age, sex and the 10 first principal components as covariates. We computed the variance explained in each cohort by rs2814778 using the following formula: 2pqβ^2^, where p is the frequency of the effect allele, q is 1-p and β is the normalized effect size of the effect allele on the HT.

To replicate the association between rs113819343, rs8090527 and platelet count, we analyzed 333 SCD patients from the Duke University Outcome Modifying Genes (OMG) cohort with whole-genome sequencing data available. We performed the association test as described above.

### Comparing effect sizes of HT-associated SNPs in SCD patients and non-SCD individuals

For each SNP-HT pair considered in the multi-ancestry PTS models, we retrieved association results from the SCD meta-analyses (above) and the published non-SCD multi-ancestry meta-analyses from the Blood-Cell Consortium.^23^ While there are 4,502 SNP-HT pairs in the PTS, we could recover results for 4,201 (93%) of them in the SCD meta-analyses. From this dataset, we corrected for multiple testing by computing a q-value for each SNP-HT association. Finally, we computed a q-value from the *P-diff* obtained (see *statistical analyses* section below).

### Association between HT or PTS with SCD-related clinical outcomes

We limited our analyses of the association between PTS and outcomes (VOC rate, ACS rate, stroke) to the large CSSCD. Further, we only considered PTS that were nominally associated with the corresponding HT (*P* < 0.05). First, we fitted Cox proportional hazard ratio models (for stroke) or quasi-Poisson regression models (for VOC and ACS rates) for the outcome on each HT (measured value), adjusting for age, sex and SCD subtype. Second, we repeated these analyses after replacing the HT by the corresponding PTS. To determine if the PTS improves model prediction beyond the measured HT, we performed an analysis of deviance. The difference between the residual deviances of the two models follows a χ2 distribution with n degrees of freedom, where *n* corresponds to the difference in the number of degrees of freedom of the two models (*i*.*e*. one degree of freedom when adding the PTS).

### Mendelian Randomization (MR)

We used a two-sample MR approach to test if HT captured by PTS causally impact SCD-related complications. We focused on the following combinations of HT and complications: WBC and neutrophil counts for survival, and HbF for VOC, ACS and stroke (for VOC and ACS, we dichotomized the data as no event vs. at least one event).

Association between SNPs and HT were performed in the GEN-MOD cohort (linear regression adjusting for sex and age), whereas association between SNPs and complications were carried out in the CSSCD (logistic regression, adjusting for sex, age and SCD subtype). For WBC and neutrophil counts, we used as instruments genome-wide significant SNPs reported by the Blood-Cell Consortium^23^ and used in the PTS analyses described above. To ensure that these variants were independent, we further pruned them using PLINK1.9b6.10 (*r*^*2*^ > 0.01 within 5-Mb windows).^31^ For HbF, we selected as instruments the six SNPs used in the PTS analyses. Although there is residual linkage disequilibrium between some of these HbF variants, we performed conditional analyses to obtain adjusted effects sizes and standard errors.

We performed all MR analyzes in RStudio (version 1.2.5033) using the TwoSampleMR package (version 0.5.5).^32^ We used the multiplicative random-effect inverse variance-weighted (IVW) approach as the main method for each MR analysis, but we also performed MR-Egger and weighted-median methods as sensitivity analyses. We assessed the validity of our statistically significant results by testing for horizontal pleiotropy (using the MR-Egger intercept test) and heterogeneity (using Cochran’s Q and I^2^ statistics).

### Statistical analyses

To compare effect sizes derived from the meta-analyses, we calculated heterogeneity *P*-values (*P-diff*) based on the following *t* statistic:^33^

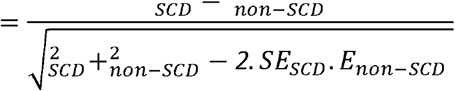

where b_SCD_ and b_non-SCD_ are the normalized effect sizes in SCD and non-SCD cohorts, respectively; SE_SCD_ and SE_non-SCD_ are the standard errors in SCD and non-SCD cohorts respectively, and *r* is the Spearman rank correlation coefficient computed using effect sizes (for the same effect allele) of all SNPs available in the meta-analyses. In our datasets, *r* ranged from -0.0016 to 0.0007 across different HT. From the *t* statistic, we can calculate a *P*-value using the normal distribution.

All statistical analyses were performed using RStudio (version 1.2.5033) or GraphPad Prism (version 9.2.0, GraphPad Software, LLC, CA).

## RESULTS

### Performance of hematological PTS in SCD patients

We investigated the phenotypic variance explained by PTS of HT in African-ancestry SCD patients from three cohorts (CSSCD, GEN-MOD, Mondor/Lyon) and in non-SCD African-American individuals from BioMe not included in the discovery effort used to generate the PTS, as well as non-SCD African-ancestry individuals from the UK Biobank. These PTS were derived from large multi-ancestry GWAS meta-analyses involving > 700,000 participants.^23^ In SCD participants, the PTS were significant (*P*-value < 0.05) in at least one cohort for nine of the 12 HT tested (**Table 1**). The non-significant PTS were for hematocrit, hemoglobin concentration and lymphocyte count. The variance explained by significant PTS ranged from 1.0 to 4.0% for RBC traits, 0.9 to 3.9% for WBC traits, and 0.6 to 4.0% for PLT traits. When we compared the performance of these PTS in SCD participants and non-SCD African-ancestry individuals, we found that all the 9 PTS with significant association explained less phenotypic variance in SCD participants (**Table 1** and **Figure 1**). One of the most striking differences was for WBC and neutrophil counts: the mean variance explained was 2.2% and 3.3% in SCD participants and 10.3% and 11.9% in non-SCD individuals, respectively (**Figure 1**).

**Table 1.** Hematological trait (HT) variance explained by polygenic trait scores (PTS) in sickle cell disease (SCD) patients. We only calculated the phenotypic variance explained for ^a^ PTS that were nominally significant (P-value < 0.05). Grey shaded cells indicate phenotypes that were not available in the corresponding study. The PTS models (except for fetal hemoglobin [HbF]) and the results from the ^b^ BioMe African American participants were reported previously^23^. For HbF, we selected six independently associated variants^26–28^. MCH: mean corpuscular hemoglobin, MCV: mean corpuscular volume, MPV: mean platelet volume.

| Phenotype | Number of variants in the PTS<sup>a</sup> | CSSCD<br>(N<sub>max</sub> = 1015) |  | GEN-MOD<br>(N<sub>max</sub> = 401) |  | Mondor/Lyon<br>(N<sub>max</sub> = 324) |  | BioME<sup>b</sup> (non-SCD)<br>(N<sub>max</sub> = 2,651) |  | UK Biobank (non-SCD)<br>(N<sub>max</sub> = 6,584) |  |
| --- | --- | --- | --- | --- | --- | --- | --- | --- | --- | --- | --- |
|  |  | Variance (%) | P-value | Variance (%) | P-value | Variance (%) | P-value | Variance (%) | P-value | Variance (%) | P-value |
| Red blood cell (RBC) traits |  |  |  |  |  |  |  |  |  |  |  |
| Hematocrit | 356 | - | 0.77 | - | 0.78 | - | 0.10 | 1.4 | 0.0071 | 2.1 | < 2.2x10<sup>-16</sup> |
| Hemoglobin | 364 | - | 1.0 | - | 0.23 | - | 0.48 | 1.3 | 7.4x10<sup>-9</sup> | 3.0 | < 2.2x10<sup>-16</sup> |
| MCH | 384 | 1.0 | 6.1x10<sup>-4</sup> | 1.3 | 0.021 | - | 0.95 | 2.2 | 5.4x10<sup>-14</sup> | 5.5 | < 2.2x10<sup>-16</sup> |
| MCV | 454 | 1.1 | 4.4x10<sup>-5</sup> | - | 0.054 | - | 0.35 | 2.0 | 9.1x10<sup>-13</sup> | 6.7 | < 2.2x10<sup>-16</sup> |
| RBC | 449 | - | 0.093 | 4.0 | 7.1x10<sup>-5</sup> | - | 0.15 | 3.8 | < 2.2x10<sup>-16</sup> | 10.6 | < 2.2x10<sup>-16</sup> |
| HbF | 6 | 17.1 | < 2.2x10<sup>-16</sup> | 22.5 | < 2.2x10<sup>-16</sup> | 26.4 | <2.2x10<sup>-16</sup> |  |  |  |  |
| White blood cell (WBC) traits |  |  |  |  |  |  |  |  |  |  |  |
| WBC | 443 | 1.2 | 7.2x10<sup>-5</sup> | 3.0 | 0.0010 | 2.2 | 0.019 | 12.2 | < 2.2x10<sup>-16</sup> | 8.3 | < 2.2x10<sup>-16</sup> |
| WBC (without chr1 - Duffy/DARC) | 389 | 0.4 | 0.013 | 3.1 | 3.8x10<sup>-4</sup> | 2.3 | 0.0067 | 1.8 | < 2.2x10<sup>-16</sup> | 1.4 | < 2.2x10<sup>-16</sup> |
| Eosinophils | 346 | 0.9 | 0.0045 | 3.4 | 2.7x10<sup>-4</sup> |  |  | 1.1 | 6.2x10<sup>-6</sup> | 3.5 | < 2.2x10<sup>-16</sup> |
| Lymphocytes | 409 | - | 0.21 | - | 0.38 |  |  | 1.4 | 7.7x10<sup>-9</sup> | 3.6 | < 2.2x10<sup>-16</sup> |
| Monocytes | 394 | 1.2 | 0.00041 | - | 0.15 |  |  | 3.9 | < 2.2x10<sup>-16</sup> | 5.1 | < 2.2x10<sup>-16</sup> |
| Neutrophils | 352 | 2.7 | 2.2x10<sup>-7</sup> | 3.9 | 2.7x10<sup>-4</sup> |  |  | 12.3 | < 2.2x10<sup>-16</sup> | 11.4 | < 2.2x10<sup>-16</sup> |
| NEU (without chr1 - Duffy/DARC) | 313 | 0.7 | 0.0059 | 3.7 | 1.4x10<sup>-4</sup> |  |  | 0.67 | 1.1x10<sup>-4</sup> | 1.2 | < 2.2x10<sup>-16</sup> |
| Platelet (PLT) traits |  |  |  |  |  |  |  |  |  |  |  |
| PLT | 553 | 0.6 | 0.0058 | 2.0 | 0.0046 | 4.0 | 2.8x10<sup>-4</sup> | 3.6 | < 2.2x10<sup>-16</sup> | 6.7 | < 2.2x10<sup>-16</sup> |
| MPV | 392 |  |  | 3.1 | 9.9x10<sup>-4</sup> |  |  | 8.6 | < 2.2x10<sup>-16</sup> | 11.8 | < 2.2x10<sup>-16</sup> |

**Figure 1.**
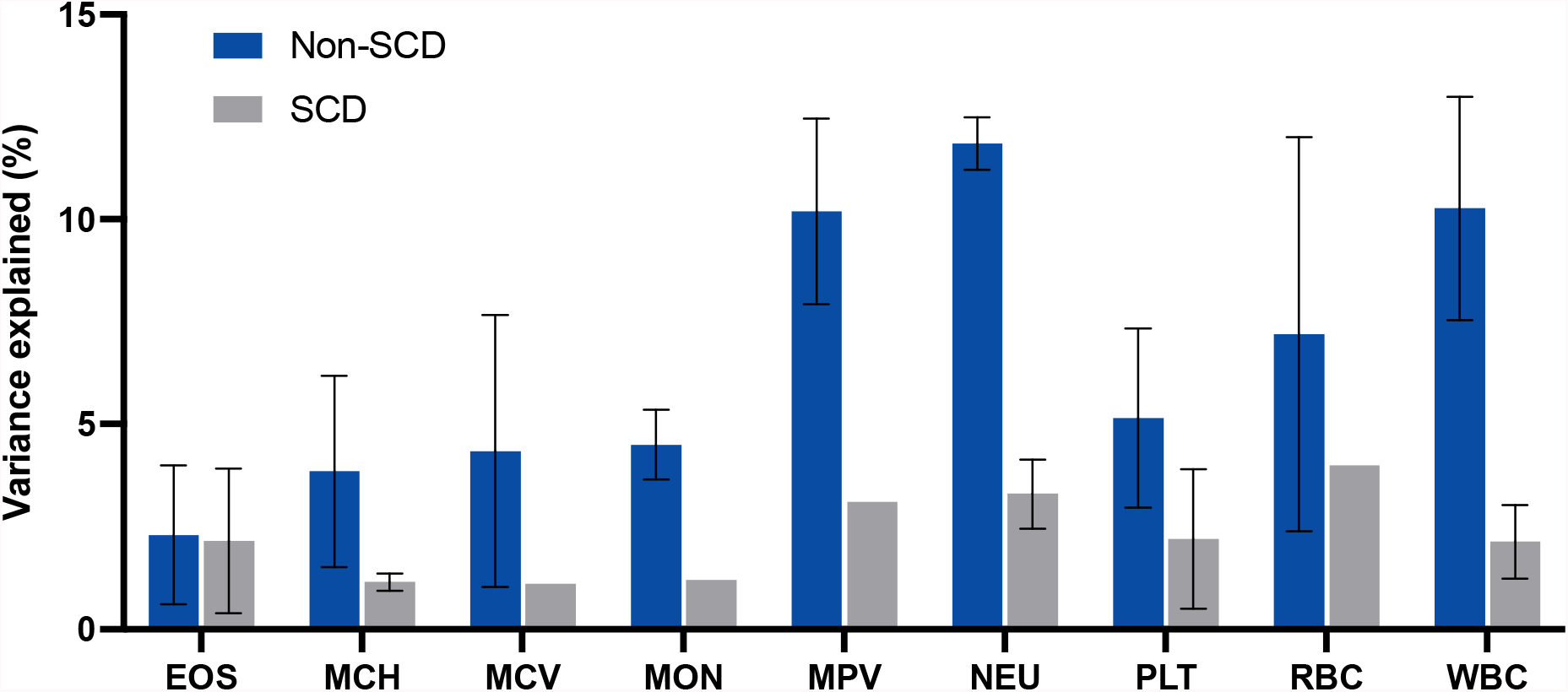
Variance explained by polygenic trait scores (PTS) for hematological traits (HT) in African-ancestry non-sickle cell disease (SCD) individuals from BioMe and the UK Biobank, and participants from three SCD cohorts (CSSCD, GEN-MOD, Mondor/Lyon). We only present the variance explained by nominally significant PTS in at least one SCD cohort (**Table 1**). When the PTS for a given HT was significant in more than one SCD cohort, we calculated the mean and standard deviation (error bars) of the variance explained. EOS: eosinophils, MCH: mean corpuscular hemoglobin, MCV: mean corpuscular volume, MON: monocytes, MPV: mean platelet volume, NEU: neutrophil count, PLT: platelet count, RBC: red blood cell count, WBC: white blood cell count.

HbF levels are an important modifier of severity in SCD. Because it is rarely measured in large non-SCD cohorts, the genetics of this trait has not been extensively studied in very large sample sizes. However, smaller GWAS in SCD patients have identified robust associations between HbF levels and genetic variants at three loci: *BCL11A, HBS1L-MYB* and the β-globin locus (reviewed in ref. ^34^). With this information, we derived an HbF PTS that includes six conditionally independent variants (**Methods** and **Table S2**). This HbF PTS was strongly associated with HbF levels in all three SCD cohorts, and explained 17.1-26.4% of the variance (**Table 1**).

### SCD partially masks the genetic effect of the Duffy/DARC null variant on WBC and neutrophil counts

To understand why the PTS under-performed in SCD patients, we carried out meta-analyses of GWAS results for the 11 HT in the three SCD cohorts, and compared effect sizes (β_SCD_) for the SNPs found in the PTS (4,201 SNP-HT pairs) with the effect sizes from multi-ancestry meta-analyses (β_non-SCD_, ref. ^23^). Across all SNPs and HT, normalized effect sizes were weakly correlated when considering the same effect alleles (Pearson’s *r* = 0.09, *P* = 2.4×10^−10^, **Figure 2A**). Among the 273 variant-HT pairs that were nominally associated in SCD meta-analyses (*P*-value < 0.05), 162 (59.3%) had a concordant direction of effect in non-SCD meta-analyses (*P* = 0.002, binomial test).

**Figure 2.**
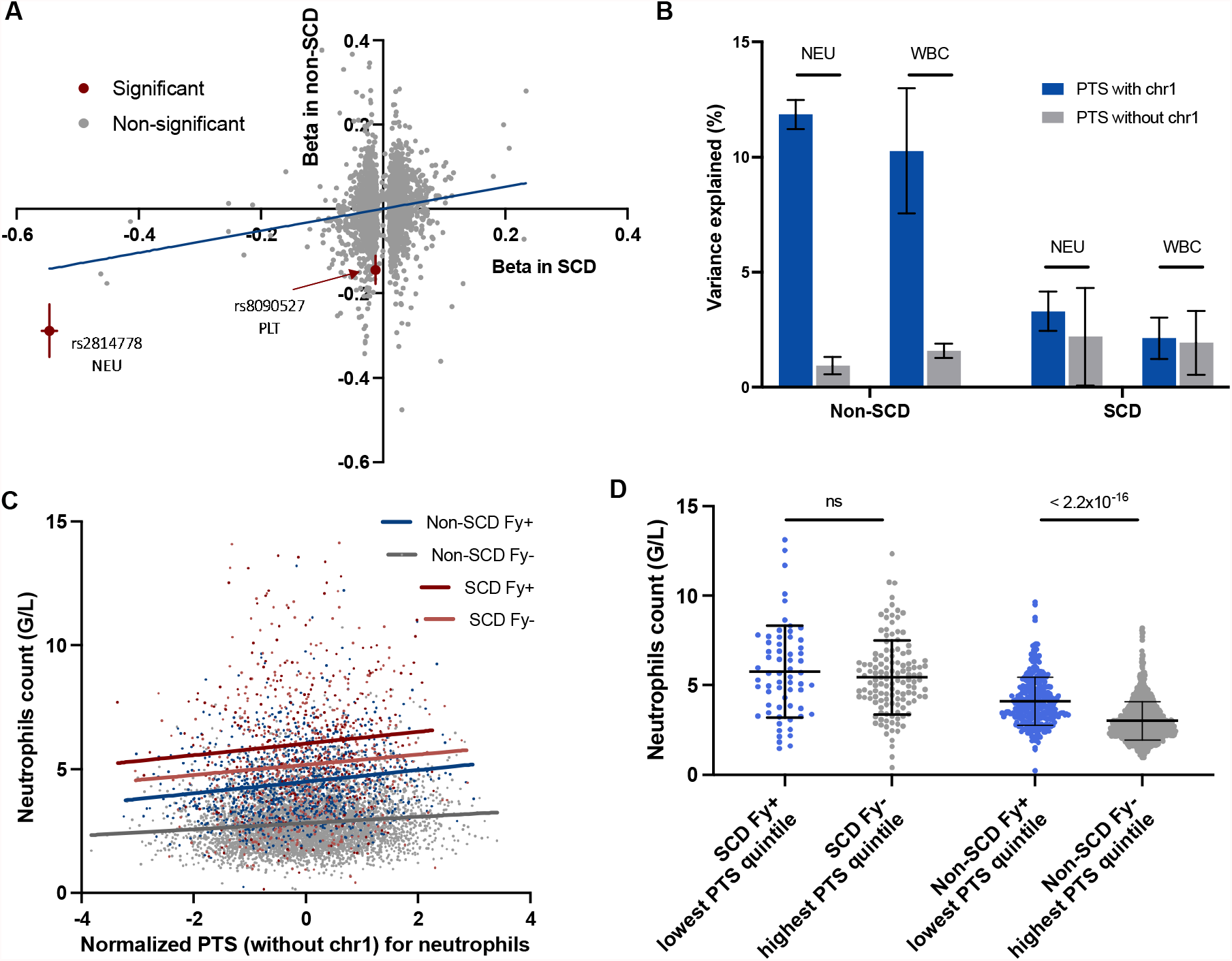
Sickle cell disease (SCD) partially masks the genetic impact of the Duffy/*DARC* null variant (rs2814778) on neutrophil count. (**A**) Comparison of effect sizes (Beta) for 3,917 SNP-hematological trait pairs in SCD patients (*x*-axis) and non-SCD participants (*y*-axis). We only considered variants with a minor allele frequency > 1% in the SCD meta-analyses. We highlight two variants with significantly different effect sizes between SCD and non-SCD individuals (see text for details). The blue line represents the best-fit linear regression line. (**B**) Variance explained (mean and standard deviation) by polygenic trait scores (PTS) for neutrophil (NEU) and white blood cell counts (WBC) with and without chromosome (chr) 1 in SCD and non-SCD individuals. (**C**) Raw neutrophil count (*y*-axis) as a function of the multi-ancestry NEU PTS (without chr 1 variants, *x*-axis) in Duffy-positive (Fy+, T/T or C/T genotypes at rs2814778) and Duffy-negative (Fy-, C/C genotype) SCD or non-SCD individuals. Regression lines for each of the four subgroups are shown. (**D**) Comparison of raw neutrophil count between Fy+ individuals with a PTS within the lowest quintile and Fy-individuals with a PTS within the highest quintile, for both SCD and non-SCD participants. ns: non-significant.

After correction for multiple testing (q-value < 0.05), we found two HT-associated variants significantly associated with HT in SCD patients, but that also had a significantly different effect size when comparing β_SCD_ and β_non-SCD_ : rs8090527 and rs2814778 (**Table S3**). We did not explore the association between the intergenic rs8090517 variant and platelet count further as we could not replicate it in an independent SCD cohort (**Table S3**). The second variant is the previously described Duffy/DARC null variant (rs2814778),^35^ and had a two-fold weaker effect on neutrophil count in SCD when compared to non-SCD individuals. Based on this observation, we wondered if the apparent poor performance of the WBC and neutrophil counts PTS in SCD participants was due to the lower impact of the Duffy/DARC null variant in this patient population. In non-SCD individuals, removing the PTS variants on chromosome 1 (to ensure that the large admixture signal due to the Duffy/DARC locus does not impact the analysis) reduced the mean variance explained for WBC from 10.3% to 1.6%, and for neutrophils from 11.9% to 0.9% (**Table 1** and **Figure 2B**). When we repeated this analysis in SCD participants, the PTS for WBC was not affected (from 2.1% to 1.9%), whereas the variance explained by the neutrophil count PTS changed slightly from 3.3% to 2.2% (**Figure 2B**).

Next, we specifically focused on the association between Duffy/*DARC* rs2814778 and WBC or neutrophil counts in SCD and non-SCD individuals. For these analyses, we used 6,627 African-ancestry participants from the UK Biobank. First, consistent with the recessive inheritance of the Duffy-negative blood group, we showed that a recessive genetic model provided a better fit with the data than the standard additive model (**Table S4**). Thus, we used a recessive model for all subsequent genetic analyses of this variant. For non-SCD individuals, the single Duffy/*DARC* variant explained 18.4-23.1% of the phenotypic variance (**Table 2**). In contrast, the associations between rs2814778 and WBC or neutrophil counts were either weak or non-significant in SCD participants, with this variant contributing only 0.9-3.3% of the variance in these HT (**Table 2**). To quantify the magnitude of the difference in effect sizes and provide meaningful clinical estimates, we calculated that the Duffy null genotype (homozygosity for the C-allele at rs2814778) was associated with a mean reduction of 0.76×10^9^ WBC/L (*P* = 0.004) and 0.84×10^9^ neutrophils/L (*P* = 1.6×10^−6^) in the CSSCD, and of 1.9×10^9^ WBC/L (*P* = 3.3×10^−164^) and 1.6×10^9^ neutrophils/L (*P* = 4×10^−199^) in the UK Biobank. When we considered both SCD status and the Duffy blood group, we found that: (1) SCD has the strongest impact on neutrophil count, (2) Duffy has a weaker effect on neutrophil count in SCD patients, and (3) the neutrophil PTS (without chromosome 1 variants) remains associated with neutrophil count in all groups (**Figure 2C**). To further illustrate how SCD modifies the effect of Duffy, we considered the neutrophil PTS (without chromosome 1 variants) quintiles and compared neutrophil count in Duffy-positive individuals with a PTS in the lowest quintile with Duffy-negative individuals with a PTS in the highest quintile (**Figure 2D**). Whereas in non-SCD UK Biobank participants Duffy outweighs the PTS effect, it is equivalent in SCD patients. Put together, our data suggest that SCD partially masks the strong effect of the *Duffy*/DARC null variant (rs2814778) on neutrophil count. Finally, we investigated whether sickle cell trait (heterozygosity for the HbS allele) also modified the Duffy/DARC effect on neutrophil count. We did not find a significant interaction between these two genotypes in the UK Biobank (P = 0.327), suggesting that the epistatic effect is specific to SCD (i.e. homozygous HbS).

**Table 2.** Genetic association results between the Duffy/DARC null variant (rs2814778) and white blood cell (WBC) and neutrophil counts. The direction of the effect is given for the CC genotype (in a recessive genetic model). Neutrophil count is not available in Mondor/Lyon. Effect sizes (Beta) and standard errors (SE) are in standard deviation units. EAF, effect allele frequency.

|  | **N** | **EAF**<br>(C-allele) | **Beta (SE)** | **P-value** | **Variance (%)** |
| --- | --- | --- | --- | --- | --- |
| *Neutrophils* |  |  |  |  |  |
| CSSCD | 934 | 0.842 | -0.353 (0.073) | $1.6 \times 10^{-6}$ | 3.3 |
| GEN-MOD | 400 | 0.939 | -0.019 (0.209) | 0.93 | - |
| UK Biobank | 6,564 | 0.906 | -1.166 (0.033) | $6.0 \times 10^{-239}$ | 23.1 |
| *White blood cells* |  |  |  |  |  |
| CSSCD | 1014 | 0.845 | -0.178 (0.064) | 0.005 | 0.9 |
| GEN-MOD | 400 | 0.939 | -0.092 (0.208) | 0.66 | - |
| Mondor/Lyon | 322 | 0.935 | -0.151 (0.214) | 0.48 | - |
| UK Biobank | 6,584 | 0.906 | -1.041 (0.036) | $5.0 \times 10^{-171}$ | 18.4 |

### GWAS of HT in SCD patients

To determine if new genetic variation could specifically modulate HT variation in SCD, we carried out GWAS for 11 blood-cell traits across all three SCD cohorts available (**Methods**). Given the relatively small sample size of the dataset, we restricted our analyses to variants with a minor allele frequency (MAF) > 1%. We found little evidence of association, except for hematocrit and hemoglobin levels (**Figure S1**). We found 24 genome-wide significant (*P* < 5×10^−8^) SNP-HT associations, including 23 at the known HbF loci and associated with hemoglobin levels, hematocrit, or RBC count (**Table S5**). The last variant, rs113819343, was associated with platelet (PLT) count in the SCD meta-analyses (*P* = 1.4×10^−8^). This variant is common in African-ancestry individuals (MAF = 5.5%, gnomAD) but rarer in European-ancestry populations (MAF = 0.098%). This variant is not associated with PLT count in the multi-ancestry (*P* = 0.72, N = 473,895) nor African-specific (*P* = 0.27, N = 15,171) meta-analyses from the Blood-Cell Consortium.^23^ Our attempt to replicate this association with PLT count in 333 SCD patients from the OMG cohort was unsuccessful (*P* = 0.69), so it is not possible to conclude if this association is real or a false positive result.

### Associations between hematological PTS and SCD-related complications

Variation in HT has been associated (prospectively) with several clinical outcomes observed in SCD patients such as death,^7^ stroke,^36^ VOC,^8^ and ACS.^9^ We could reproduce most of those results in the genotyped subset of the CSSCD (**Table S6**). Having established that multi-ancestry PTS are associated with HT in SCD participants (although to a lesser extent than in non-SCD individuals), we asked if they were also associated with these complications in the CSSCD. For these analyses, we used nominally significant PTS (**Table 1**) for which the corresponding HT was also significantly associated with the complication (**Table S6**). In total, we tested six PTS-complication combinations, and found significant associations for PTS_HbF_-stroke and PTS_HbF_-ACS (**Table 3**).

**Table 3.** The polygenic trait score (PTS) for fetal hemoglobin (HbF) levels is associated with stroke and acute chest syndrome (ACS), and improves the prediction of vaso-occlusive crises (VOC) rates. We carried out these analyses in 1,278 genotyped CSSCD participants. To compare predictive models, we performed an analysis of deviance and compared a baseline model (HT, age, sex, α-thalassemia) with a model that included the same predictors as well as the PTS (**Methods**).

| Outcome | PTS | Association of the PTS with the outcome |  | Comparison of predictive models with and without PTS |  |
| --- | --- | --- | --- | --- | --- |
| | | HR [95% CI] or beta (SE) | P-value | $\chi^2$ | P-value |
| Stroke (N<sub>cases</sub> = 104, N<sub>controls</sub> = 1,168) | HbF | 0.75 [0.61-0.91] | 0.004 | 0.14 | 0.70 |
|  | WBC | 1.01 [0.84-1.22] | 0.92 | 2.07 | 0.15 |
|  | NEU | 0.98 [0.81-1.19] | 0.86 | 3.21 | 0.07 |
| ACS rate (N = 1,271) | HbF | -0.21 (0.06) | 0.0003 | 0.231 | 0.63 |
|  | EOS | -0.02 (0.05) | 0.78 | 0.68 | 0.41 |
| VOC rate (N = 1,271) | HbF | 0.05 (0.05) | 0.26 | 14.2 | 0.0002 |

We extended these analyses to determine if the PTS could improve the predictive performance of the models beyond the baseline HT measures. We reasoned that because the PTS capture HT heritable variation, they would more faithfully represent “life-long exposure” and add information that is independent from HT lab measure imprecisions. The models did not improve for stroke and ACS rate, but we found that PTS_HbF_ improved the prediction of VOC, consistent with previous reports (**Table 3**).^37,38^ Interestingly, additional analyses revealed that PTS_HbF_ improved the prediction of VOC in patients with low HbF lab values (< 10%), levels at which HbF is not associated with VOC (**Table S7**). For high HbF values (> 10%), HbF is strongly associated with VOC and adding PTS_HbF_ did not improve the predictive model.

### Quantifying the causal impact of HbF levels on SCD complications by MR

Finally, we sought to confirm some of the causal effects of HT on SCD complications as reported in the literature: the protection that high HbF levels confer against stroke,^39,40^ ACS^9^ and VOC, ^8^ as well as the increased risk of death associated with high WBC^7^ and neutrophil counts.^41^ To this end, we performed two-sample MR using the pseudo-independent SNPs from the PTS of HbF, WBC and neutrophils (**Methods**). We found a causal association between HbF and stroke: a one standard deviation increase in genetically-determined HbF levels (corresponding to 4.8% of HbF) decreases by 36% the risk of stroke (odds ratio [95% confidence interval] = 0.64 [0.49-0.83], *P* = 0.0008, **Figure 3**). Although the direction of the effect was similar using MR-Egger and weighted median, these analyzes were not statistically significant, suggesting insufficient power (**Table S8**). We found no heterogeneity in the effect (I^2^ = 0%) and confirmed the absence of horizontal pleiotropy (Egger intercept, -0.07, *P* = 0.75). All other MR tests were non-significant (**Table S8**).

**Figure 3.**
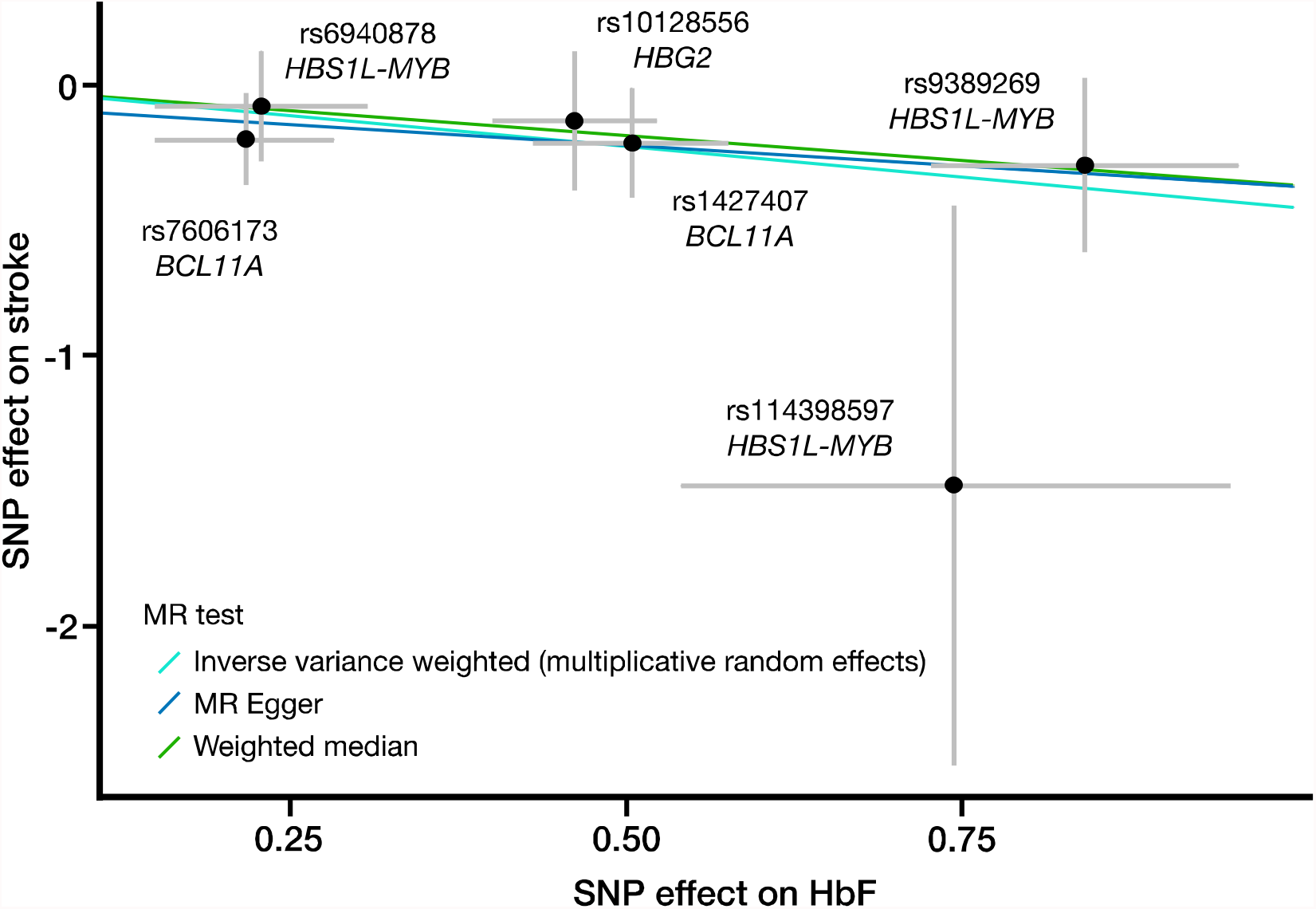
Mendelian randomization (MR) results for fetal hemoglobin (HbF) levels on stroke. Each dot represents one of the HbF-associated SNP, with its corresponding effect on normalized HbF levels (*x*-axis, standard deviation units) and stroke risk (*y*-axis, logistic regression beta). The analysis without rs114398597 is significant; see **Table S8** for details.

## DISCUSSION

In this study, we showed that PTS for HT derived in non-SCD individuals largely underperformed when tested in SCD patients. We hypothesize that this is due to an epistatic effect of the SCD genotype on the large number of variants that contribute to polygenic variation in HT. In particular, we found a dramatic impact of SCD on the magnitude of the effect of the Duffy/*DARC* null rs2814778 variant on neutrophil count. Potentially because the phenotypic variance explained by these PTS for HT remain low, we could not detect an effect of these PTS on disease expressivity when considering different SCD-related complications. However, a small set of six HbF-associated variants was useful to predict VOC when considered as a PTS, and to quantify the causal protective impact of HbF increase on stroke risk reduction.

Several non-mutually exclusive factors could explain why PTS for HT were not as predictive in SCD patients. These patients have different baseline levels and distribution values for several HT such as hemoglobin and WBC counts. This is in part due to the direct hemolytic effect of hemoglobin S, but also to the broad consequences of SCD such as the induction of a chronic inflammatory state that can lead to cytokines-driven higher WBC count.^42,43^ Moreover, the frequent intercurrent complications (*e*.*g*. VOC) experienced throughout SCD natural history could result in greater variability in HT values. Finally, SNP genotyping arrays do not capture all structural variants, which are the main alterations in α-thalassemia, a major determinant of mean corpuscular hemoglobin and mean corpuscular volume. Thus, the performance of HT PTS will improve once comprehensive whole-genome sequencing of SCD cohorts becomes available.

The C-allele of the Duffy/*DARC* null variant (rs2814778) results in erythroid-specific loss of expression of the Duffy/*DARC* (also known as *ACKR1*) chemokine reservoir expression gene.^44,45^ It has been known that the Duffy/*DARC* null variant is strongly associated with lower circulating neutrophil and WBC counts in SCD and non-SCD individuals due to extravasation to tissues,^46–50^ and in particular the spleen.^51^ Although it is unclear why SCD partially masks the effect of this variant on neutrophil count, we may speculate that the precocious and pervasive splenic atrophy observed in SCD patients could lead to a reduced reservoir size. Additionally, various Duffy/*DARC* variants in non-SCD and SCD individuals have been shown to affect the binding of DARC with inflammatory markers such as interleukin 8.^52–54^ Definitive data are still lacking, but the proinflammatory state of SCD patients may contribute to the discrepancy in effect observed.^55^

Whether or not the Duffy phenotype is associated with complications in SCD patients remains unclear.^56–61^ The potential consequences of the Duffy/*DARC* status in SCD would be linked to circulating proinflammatory cytokines and especially the resulting effect on neutrophil and WBC counts, which has been implicated in several SCD complications.^7,12,13^ However, our data showed that the Duffy/*DARC* negative phenotype is not a good proxy for neutrophil count in SCD patients in contrast to non-SCD individuals in whom it accounts for a large fraction of the phenotypic variance. Thus, while it remains possible that neutrophils causally contribute to SCD clinical heterogeneity, we have not been able to use genetics, including MR, to confirm this epidemiological observation owing to the relatively weak effect of the neutrophil-associated variants in SCD patients.

Although complex multi-ancestry PTS for general HT that included 100s of variants performed poorly in SCD patients, we showed that a simple PTS for HbF made of six variants at three loci can capture a large fraction of the phenotypic variance in this important SCD modifier. One major difference between the general HT and HbF is that we developed PTS_HbF_ using GWAS data from SCD patients. Interestingly, we observed an association between PTS_HbF_ and ACS or stroke in the large CSSCD. We validated previous reports that a PTS for HbF can improve the prediction of VOC,^37,38^ and further discovered that this PTS_HbF_ was useful in the subset of patients with HbF levels < 10%. Our limited sample size prevented powerful analyses of causality, yet we could use MR to quantify the protective effect of HbF on stroke risk in SCD patients, although we acknowledge that the Winner’s curse could have biased upward our estimate. This result is consistent with clinical trials that have shown that the HbF-inducing drug hydroxyurea (HU) can reduce stroke risk in primary prevention in SCD.^62,63^ Furthermore, although HU can improve SCD through different mechanisms,^64^ our MR analysis is useful to partition its effect and specifically quantify how genetically-determined (lifelong) HbF levels modulate stroke risk. Given its hematological rationale, simplicity, and performance, we believe that the time is right to formally test in a pragmatic randomized clinical trial whether PTS_HbF_ is useful to manage clinical heterogeneity in SCD patients.

Our findings have implications beyond SCD. While PTS have been shown to modulate the penetrance of monogenic mutations in diseases such as coronary artery disease (CAD) and familial breast and colorectal cancers,^65^ much less is known about their effect on expressivity (or disease severity).^4^ This distinction is important because, although they may not cause the disease, several clinical variables and other endophenotypes that are captured by PTS can strongly modify disease severity (*e*.*g*. PTS for kidney functions in the context of hypertension or CAD, PTS for retinopathy/cataract in diabetic patients). Our analyses indicate that simply translating the genetics of polygenic traits learned in healthy individuals to diseased populations may not provide the expected gain in risk stratification in the context of precision medicine. Fortunately, large biobanks and other cohorts should soon enable powerful GWAS for genetic modifiers in > 10,000 patients who all suffer from the same disease.

## Data Availability

The CSSCD genetic dataset is available on the database of Genotypes and Phenotypes (dbGaP: https://www.ncbi.nlm.nih.gov/gap/), accession phs000366.v1.p1. The GEN-MOD, Mondor/Lyon and BioMe data are available upon requests to the authors. The UK Biobank dataset is publicly available (https://www.ukbiobank.ac.uk/).

## SUPPLEMENTAL DATA

Supplemental data include one figure and eight tables.

## ACKNOWLEDGMENTS

We thank all participants for their contribution to this project. We thank Gabrielle Boucher for statistical support. T.P. is a recipient of a Charles Bruneau Foundation fellowship award and merit scholarship program for foreign students from the Ministry of Education and Higher Education of Quebec. This work was funded by the Canadian Institutes of Health Research (PJT #156248), Bioverativ, a Sanofi Company, and the Canada Research Chair Program (to G.L.). GEN-MOD samples and data collection were supported by NIH grant HL-68922. A.A-K., M.J.T. and establishment and analysis of the OMG cohort have been funded by NHLBI (R01HL68959, HL79915, HL70769, HL87681). This research has been conducted using the UK Biobank Resource under Application Number 11707.

## DATA AND CODE AVAILABILITY

The CSSCD genetic dataset is available on the database of Genotypes and Phenotypes (dbGaP: https://www.ncbi.nlm.nih.gov/gap/), accession phs000366.v1.p1. The UK Biobank dataset is publicly available (https://www.ukbiobank.ac.uk/). The GEN-MOD, Mondor/Lyon and BioMe datasets and code supporting the current study have not been deposited in a public repository because data are not public but are available from the corresponding author on request.

## AUTHOR CONTRIBUTIONS

T.P. and G.L. designed the study. T.P., K.S.L. and M.E.G. performed the analyses. T.P., A.L.P.H.A.O., M.E.G, C.B., A.E.A.-K., M.J.T., F.G., P.J., P.B and G.L. collected the clinical and genetic data. T.P. and G.L. drafted the paper. G.L. supervised the study. All authors participated in data interpretation, revised the manuscript for critical content and approved the final manuscript.

