## Supplemental data for "Variation and impact of polygenic hematological traits in monogenic sickle cell disease"

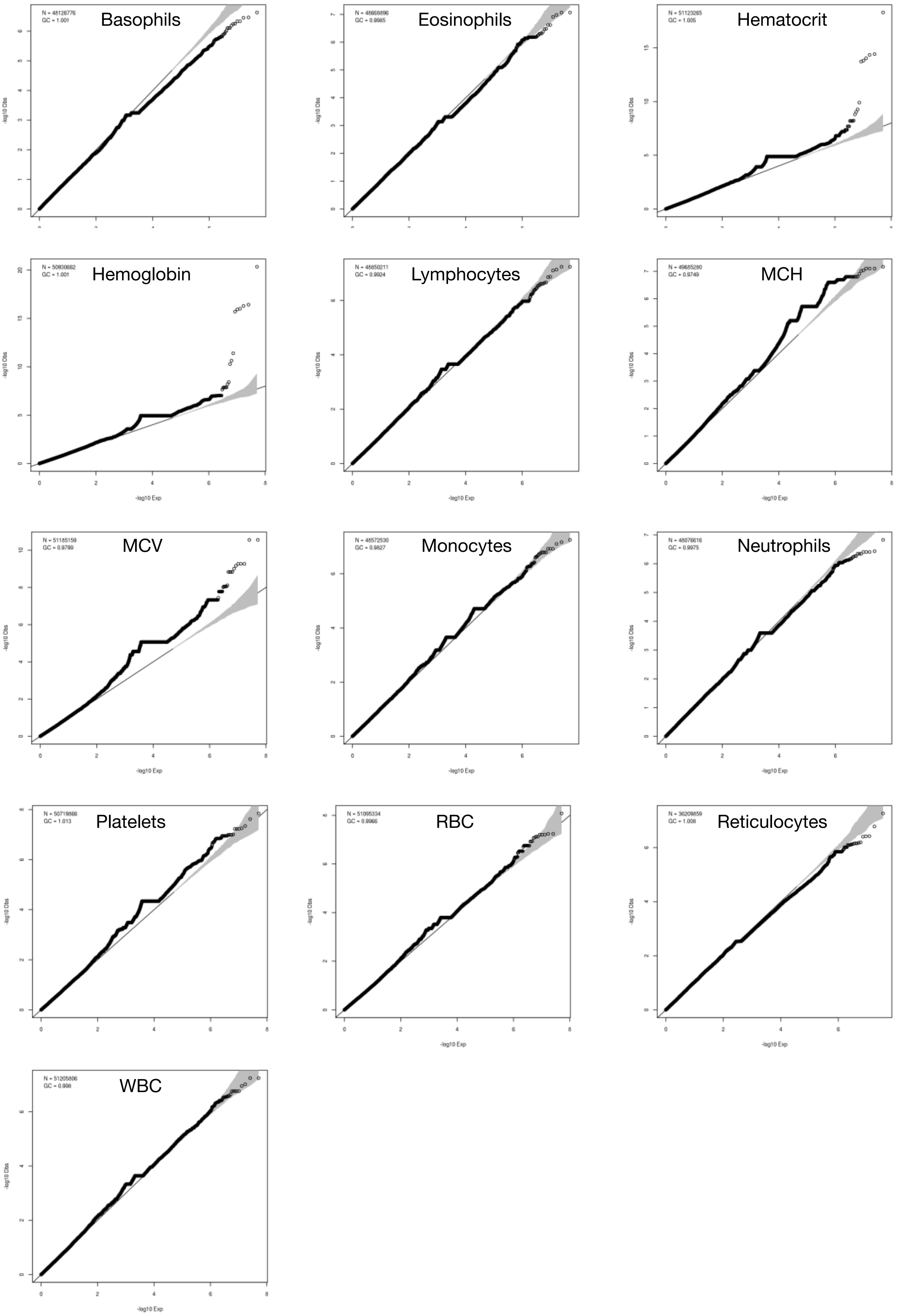


**Figure S1.** Quantile-quantile plots of the meta-analyses of the GWAS results for hematological traits performed in sickle cell disease patients. λgc: lambda genomic control, inflation factor.

**Table S1.** Population demographics. Alpha-thalassemia status is not available for the GEN-MOD and Mondor/Lyon cohorts. In the CSSCD cohort, vaso-occlusive crises (VOC) are defined as painful episodes requiring emergency room visits. For VOC and acute chest syndrome (ACS), rates are defined as the number of episodes per year. NA, not available.

| Characteristic | CSSCD | GEN-MOD | Mondor/Lyon | UK Biobank (African ancestry) |
|---|---|---|---|---|
| Number of individuals included | 1,278 | 406 | 372 | 6,627 |
| Sex, male/female | 616/662 | 222/184 | 139/233 | 2,930/3,697 |
| Age (year), mean ± SD | 14 ± 12 | 31 ± 9 | 35 ± 13 | 52 ± 8 |
| Genotypes, HbSS/HbSS α-thal | 883/395 | 406/NA | 372/NA | NA |
| VOC rate, mean ± SD | 0.81 ± 1.42 | NA | NA | NA |
| ACS rate, mean ± SD | 0.13 ± 0.29 | NA | NA | NA |
| Stroke (%) | 105 (8.3) | NA | NA | NA |
| Death (%) | 44 (3.4) | 19 (4.7) | NA | NA |
| Hematocrit (%), mean ± SD | 24.79 ± 3.96 | 25.7 ± 4.53 | 26.05 ± 4.24 | 40.3 ± 3.85 |
| Hb (g/L), mean ± SD | 8.44 ± 1.25 | 8.76 ± 1.32 | 8.786 ± 1.27 | 13.61 ± 1.35 |
| MCH (pg), mean ± SD | 30.04 ± 2.88 | 29.30 ± 4.14 | 29.25 ± 4.93 | 29.66 ± 2.44 |
| MCV (fL), mean ± SD | 89.12 ± 8.57 | 86.90 ± 10.19 | 85.32 ± 10.81 | 87.80 ± 6.13 |
| RBC ($10^9$/L), mean ± SD | 2.80 ± 0.56 | 3.00 ± 0.78 | 3.367 ± 4.68 | 4.61 ± 0.51 |
| HbF (%), mean ± SD | 6.45 ± 4.37 | 6.66 ± 4.81 | 7.973 ± 6.37 | NA |
| WBC ($10^6$/L), mean ± SD | 12.19 ± 3.78 | 10.60 ± 3.67 | 10.62 ± 3.01 | 5.81 ± 1.66 |
| Eosinophils ($10^6$/L), mean ± SD | 0.43 ± 0.42 | 0.27 ± 0.31 | NA | 0.16 ± 0.14 |
| Basophils ($10^6$/L), mean ± SD | 0.06 ± 0.10 | 0.09 ± 0.01 | NA | 0.03 ± 0.03 |
| Lymphocytes ($10^6$/L), mean ± SD | 4.78 ± 2.44 | 3.59 ± 1.46 | NA | 2.12 ± 0.67 |
| Monocytes ($10^6$/L), mean ± SD | 0.88 ± 0.56 | 0.81 ± 0.49 | NA | 0.39 ± 0.16 |
| Neutrophils ($10^6$/L), mean ± SD | 5.42 ± 2.44 | 5.84 ± 2.67 | NA | 3.10 ± 1.27 |
| Platelets ($10^9$/L), mean ± SD | 449.2 ± 147.00 | 391.0 ± 120.3 | 382.3 ± 131.44 | 253.43 ± 59.34 |
| MPV (fL), mean ± SD | NA | 8.62 ± 1.02 | NA | 9.68 ± 1.17 |

**Table S2**. SNPs selected to derive the fetal hemoglobin (HbF) polygenic trait score (PTS). We selected six independently HbF-associated variants and used the published effect sizes (normalized HbF betas conditioned on all other HbF variants) as weights in an additive PTS model.

| SNP coordinate (hg38) | Effect allele | Effect size | SNP ID | Gene | Reference |
|---|---|---|---|---|---|
| chr2:60490908_G_T | T | 0.6634 | rs1427407 | *BCL11A* | [27] |
| chr2:60498316_C_G | C | -0.2632 | rs7606173 | *BCL11A* | [27] |
| chr6:135045171_A_G | G | -0.2342 | rs6940878 | *HBSL1-MYB* | [28] |
| chr6:135106021_C_T | C | 0.581725 | rs9389269 | *HBSL1-MYB* | [28] |
| chr6:135107536_A_G | G | 0.677248 | rs114398597 | *HBSL1-MYB* | [28] |
| chr11:5242453_C_T | T | 0.421 | rs10128556 | ß-globin locus | [26] |

**Table S3**. SNPs with a significantly different effect size on hematological trait (HT) between sickle cell disease (SCD) and non-SCD individuals. For all the 4201 SNP-HT pairs present in the 11 PTS, we compared the meta-analyzed effect sizes between SCD and non-SCD datasets using the heterogeneity *t* statistic (see **Methods**). We found only two SNPs with both a significant difference in the effect size and a significant association with HT in the SCD meta-analysis after correction for multiple testing (q-value < 0.05). Note that for the Duffy/*DARC* variant (rs2814778), the effect allele frequency (EAF) is very different between the SCD cohorts (C-allele, 85%) and the multi-ancestry meta-analyses (C-allele, 2%) because ~98% of the samples were of non-African ancestry. SE: standard error. The association between rs8090527 and PLT count could not be replicated in 333 SCD participants from the OMG cohort (C-allele frequency = 0.34 ; Beta_C_allele = 0.0961 ; SE = 0.0842 ; P-value = 0.25).

| | | | | **SCD (this study)** | | | | **Non-SCD**[23] | | | **Heterogeneity ($\beta_{SCD} = \beta_{non\text{-}SCD}$?)** | |
|---|---|---|---|---|---|---|---|---|---|---|---|---|
| **HT** | **Position (hg38)** | **SNP** | **Ref/Effect allele** | **EAF** | **Beta (SE) SCD** | **P-value** | **q-value** | **EAF** | **Beta (SE)** | **P-value** | **P-value t stat** | **q-value t stat** |
| NEU | 1:159204893 | rs2814778 | T/C | 0.85 | -0.288 (0.063) | 4.1E-6 | 0.015 | 0.02 | -0.546 (0.012) | 0 | 4.1E-6 | 0.031 |
| PLT | 18:51257657 | rs8090527 | T/C | 0.37 | -0.144 (0.034) | 1.9E-5 | 0.036 | 0.51 | -0.012 (0.002) | 1.1E-13 | 1.9E-5 | 0.037 |

**Table S4.** Comparison of the Duffy/*DARC* null rs2814778 effect on neutrophil and white blood cell (WBC) counts between the additive and recessive genetic models. We computed the effect of rs2814779 on normalized neutrophil and WBC counts (after adjusting for age and sex) using both additive and recessive model. We used the first 10 principal components as covariates. In the UK Biobank, we only analyzed participants of African ancestry. The effect size (Beta and standard error [SE]) is for the C-allele (additive model) or the CC genotype (recessive model). We calculated the phenotypic variance explained only for nominally significant associations. Neutrophil count is not available in the Mondor/Lyon cohort. EAF: effect allele frequency, N: sample size, SE: standard error.

| | | | **Additive** | | | **Recessive** | | |
|---|---|---|---|---|---|---|---|---|
| **Cohort** | **N** | **EAF** | **Beta (SE)** | **P-value** | **Variance (%)** | **Beta (SE)** | **P-value** | **Variance (%)** |
| *Neutrophil count* | | | | | | | | |
| CSSCD | 934 | 0.842 | -0.313 (0.065) | $1.58 \times 10^{-6}$ | 2.61 | -0.353 (0.073) | $1.64 \times 10^{-6}$ | 3.34 |
| GEN-MOD | 400 | 0.939 | -0.083 (0.19) | 0.661 | - | -0.019 (0.209) | 0.927 | - |
| UK Biobank | 6564 | 0.906 | -0.986 (0.03) | $2.03 \times 10^{-210}$ | 16.51 | -1.166 (0.033) | $5.96 \times 10^{-239}$ | 23.12 |
| *White blood cell count* | | | | | | | | |
| CSSCD | 1014 | 0.845 | -0.164 (0.056) | 0.004 | 0.71 | -0.178 (0.064) | 0.005 | 0.84 |
| GEN-MOD | 400 | 0.939 | -0.135 (0.189) | 0.474 | - | -0.092 (0.208) | 0.657 | - |
| Mondor/ Lyon | 322 | 0.935 | -0.053 (0.187) | 0.775 | - | -0.151 (0.214) | 0.481 | - |
| UK Biobank | 6584 | 0.906 | -0.879 (0.032) | $1.56 \times 10^{-150}$ | 13.13 | -1.041 (0.036) | $4.96 \times 10^{-171}$ | 18.42 |

**Table S5.** Results of the GWAS meta-analyses for hematological traits (HT) in sickle cell disease patients. We only report variants that reached genome-wide significance ($P < 5x10^{-8}$). Except for the variant on chromosome (chr) 3 associated with platelet (PLT) count, all other variants associate with red blood cell (RBC) traits and are at the *BCL11A* (chr 2) and *HBS1L-MYB* (chr 6) loci. EAF: effect allele frequency, HCT: hematocrit, HGB: hemoglobin.

| HT | Chr | Position (hg38) | Ref | Alt | EAF | Beta | Standard error | P-value | Direction | $I^2$ | Hetero. P-value |
|---|---|---|---|---|---|---|---|---|---|---|---|
| HCT | 2 | 60490908 | T | G | 0.2689 | 0.303 | 0.034 | 5.09E-19 | +++ | 0 | 0.9612 |
| HGB | 2 | 60490908 | T | G | 0.2697 | 0.3187 | 0.0338 | 4.47E-21 | +++ | 0 | 0.8823 |
| RBC | 2 | 60490908 | T | G | 0.2686 | 0.1918 | 0.0333 | 8.30E-09 | +++ | 28.8 | 0.2453 |
| HCT | 2 | 60492835 | C | A | 0.2935 | 0.2512 | 0.0327 | 1.54E-14 | +++ | 0 | 0.8708 |
| HGB | 2 | 60492835 | C | A | 0.2935 | 0.2707 | 0.0326 | 9.93E-17 | +++ | 0 | 0.7614 |
| HCT | 2 | 60493816 | A | G | 0.2933 | 0.2504 | 0.0327 | 1.88E-14 | +++ | 0 | 0.8406 |
| HGB | 2 | 60493816 | A | G | 0.2932 | 0.27 | 0.0326 | 1.17E-16 | +++ | 0 | 0.7394 |
| HCT | 2 | 60494905 | T | C | 0.3856 | -0.1801 | 0.031 | 5.94E-09 | --- | 0 | 0.5633 |
| HGB | 2 | 60494905 | T | C | 0.3844 | -0.1795 | 0.0309 | 6.11E-09 | --- | 0 | 0.4842 |
| HCT | 2 | 60495961 | C | CA | 0.2842 | 0.2561 | 0.033 | 9.32E-15 | +++ | 0 | 0.836 |
| HGB | 2 | 60495961 | C | CA | 0.2846 | 0.2705 | 0.0329 | 1.95E-16 | +++ | 0 | 0.7371 |
| HCT | 2 | 60496951 | T | C | 0.2891 | 0.258 | 0.0329 | 4.53E-15 | +++ | 0 | 0.8085 |
| HGB | 2 | 60496951 | T | C | 0.2895 | 0.2746 | 0.0327 | 5.05E-17 | +++ | 0 | 0.6949 |
| HCT | 2 | 60496952 | G | T | 0.2902 | 0.2583 | 0.0329 | 3.82E-15 | +++ | 0 | 0.8844 |
| HGB | 2 | 60496952 | G | T | 0.2903 | 0.2756 | 0.0327 | 3.73E-17 | +++ | 0 | 0.735 |
| HCT | 2 | 60498316 | C | G | 0.4224 | -0.183 | 0.0303 | 1.58E-09 | --- | 0 | 0.7167 |
| HGB | 2 | 60498316 | C | G | 0.4213 | -0.178 | 0.0302 | 3.79E-09 | --- | 0 | 0.5893 |
| PLT | 3 | 123967670 | C | T | 0.0566 | -0.4085 | 0.072 | 1.42E-08 | --- | 49.3 | 0.1389 |
| HCT | 6 | 135078218 | A | G | 0.0267 | 0.5813 | 0.095 | 9.25E-10 | +++ | 46 | 0.1572 |
| HGB | 6 | 135078218 | A | G | 0.0269 | 0.6205 | 0.0945 | 5.24E-11 | +++ | 66.8 | 0.04897 |
| HCT | 6 | 135086355 | G | A | 0.0287 | 0.5892 | 0.0916 | 1.25E-10 | +++ | 41.9 | 0.1789 |
| HGB | 6 | 135086355 | G | A | 0.0288 | 0.6324 | 0.0912 | 3.99E-12 | +++ | 65 | 0.05735 |
| HCT | 6 | 135097526 | C | G | 0.0281 | 0.5744 | 0.0925 | 5.35E-10 | +++ | 46.5 | 0.1544 |
| HGB | 6 | 135097526 | C | G | 0.0282 | 0.6157 | 0.0921 | 2.28E-11 | +++ | 68.5 | 0.04185 |

**Table S6.** Association between sickle cell disease (SCD)-related complications and hematological traits (HT) or corresponding polygenic trait scores (PTS) in 1,278 genotyped participants from the CSSCD. We used Cox proportional-hazards models for stroke, and quasi-Poisson regression for acute chest syndrome (ACS) and vaso-occlusive crises (VOC) rates. We first tested the association between raw HT and complications, correcting for age at recruitment, sex, and α-thalassemia status. We then replaced the HT by its corresponding normalized PTS (using inverse normal transformation). EOS: eosinophils, LYM: lymphocytes, MCH: mean corpuscular hemoglobin, MCV: mean corpuscular volume, MON: monocytes, MPV: mean platelet volume, NEU: neutrophil count, PLT: platelet count, WBC: white blood cell count, SE: standard error.

| | **Hematological trait** | | | **Normalized PTS for hematological trait** | | |
|---|---|---|---|---|---|---|
| | Effect | SE | P-value | Effect | SE | P-value |
| *Stroke, Cox proportional-hazards model* | | | | | | |
| MCH | 0.03 | 0.05 | 0.58709 | 0.00 | 0.10 | 0.9742 |
| MCV | 0.02 | 0.02 | 0.13100 | 0.02 | 0.10 | 0.8649 |
| HbF | -0.11 | 0.03 | 0.00088 | -0.29 | 0.10 | 0.0040 |
| WBC | 0.09 | 0.03 | 0.00118 | 0.01 | 0.10 | 0.9223 |
| EOS | 0.05 | 0.27 | 0.86110 | -0.17 | 0.10 | 0.0855 |
| LYM | 0.09 | 0.05 | 0.05720 | 0.02 | 0.10 | 0.8706 |
| MONO | 0.31 | 0.19 | 0.09800 | -0.11 | 0.10 | 0.2833 |
| NEU | 0.11 | 0.04 | 0.01320 | -0.02 | 0.10 | 0.8623 |
| PLT | 0.00 | 0.00 | 0.61710 | -0.10 | 0.10 | 0.3091 |
| *ACS, quasi-Poisson regression* | | | | | | |
| MCH | -0.03 | 0.02 | 0.14400 | 0.00 | 0.06 | 0.9340 |
| MCV | -0.01 | 0.01 | 0.13200 | 0.01 | 0.06 | 0.8200 |
| HbF | -0.07 | 0.02 | 0.00001 | -0.21 | 0.06 | 0.0003 |

| | | | | | | |
|---|---|---|---|---|---|---|
| WBC | 0.03 | 0.01 | 0.07360 | -0.02 | 0.05 | 0.7200 |
| EOS | 0.24 | 0.12 | 0.04920 | -0.02 | 0.05 | 0.7790 |
| LYM | 0.04 | 0.02 | 0.07530 | -0.04 | 0.05 | 0.4090 |
| MONO | 0.02 | 0.10 | 0.81500 | -0.05 | 0.05 | 0.3200 |
| NEU | 0.03 | 0.02 | 0.25200 | 0.00 | 0.05 | 0.9320 |
| PLT | 0.00 | 0.00 | 0.57800 | 0.00 | 0.05 | 0.9400 |
| *VOC, quasi-Poisson regression* | | | | | | |
| MCH | -0.04 | 0.02 | 0.03910 | 0.03 | 0.05 | 0.5716 |
| MCV | -0.01 | 0.01 | 0.21700 | 0.09 | 0.05 | 0.0405 |
| HbF | -0.03 | 0.01 | 0.04250 | 0.05 | 0.05 | 0.2537 |
| WBC | -0.01 | 0.01 | 0.51092 | 0.00 | 0.05 | 0.9688 |
| EOS | -0.04 | 0.13 | 0.76674 | 0.04 | 0.05 | 0.4519 |
| LYM | -0.04 | 0.03 | 0.10154 | 0.01 | 0.05 | 0.7671 |
| MONO | 0.03 | 0.10 | 0.72143 | 0.05 | 0.05 | 0.2987 |
| NEU | 0.01 | 0.02 | 0.77612 | 0.03 | 0.05 | 0.5965 |
| PLT | 0.00 | 0.00 | 0.24469 | 0.02 | 0.05 | 0.7127 |

**Table S7.** The polygenic trait score (PTS) for fetal hemoglobin (HbF) levels improves the prediction of vaso-occlusive crises (VOC) rates for patients with low (< 10%) HbF levels. We carried out these analyses in 1,139 CSSCD participants. To compare the predictive models, we performed an analysis of deviance and compared a baseline model (HT, age, sex, α-thalassemia) with a model that included the same predictors as well as $PTS_{HbF}$ (**Methods**).

| | **HbF < 10% (n = 930)** | | **HbF ≥ 10% (n = 209)** | |
|---|---|---|---|---|
| | **Beta (SE)** | **P-value** | **Beta (SE)** | **P-value** |
| *Baseline model* (VOC prediction without $PTS_{HbF}$) | | | | |
| HbF | -0.002 (0.022) | 0.93 | -0.14 (0.05) | 0.0027 |
| Age | 0.017 (0.004) | $7.4x10^{-5}$ | 0.029 (0.008) | 0.00015 |
| Sex | 0.003 (0.11) | 0.98 | -0.039 (0.23) | 0.87 |
| α-thalassemia | 0.03 (0.03) | 0.23 | -0.002 (0.06) | 0.98 |
| *Complete model* (VOC prediction with $PTS_{HbF}$) | | | | |
| HbF | -0.019 (0.065) | 0.44 | -0.15 (0.12) | 0.0024 |
| Age | 0.016 (0.004) | 0.0002 | 0.027 (0.008) | 0.0007 |
| Sex | 0.011 (0.110) | 0.92 | -0.037 (0.23) | 0.87 |
| α-thalassemia | 0.035 (0.028) | 0.21 | -0.004 (0.06) | 0.95 |
| $PTS_{HbF}$ | 0.110 (0.065) | 0.09 | 0.15 (0.12) | 0.23 |
| *Comparison between the two models* | | | | |
| | **χ2** | **P-value** | **χ2** | **P-value** |
| Residual deviance difference | 52.1 | $5.3x10^{-13}$ | 2.39 | 0.12 |

**Table S8.** Mendelian randomization (MR) results for hematological traits (HT) and sickle cell disease (SCD)-related complications. We selected the SNPs included in the polygenic trait scores as instruments. We used a two-samples MR approach to test the causality of several HT on SCD complication: white blood cells (WBC) and neutrophil counts with death, and fetal hemoglobin (HbF) with acute chest syndrome (ACS), stroke and vaso-occlusive crises (VOC).

| Outcome | Exposure (HT) | Method | Number of SNPs | Beta | Standard error | P-value |
|---|---|---|---|---|---|---|
| Stroke | HbF | Inverse variance weighted (multiplicative random effects) | 6 | -0.452 | 0.135 | 8.27E-04 |
| | | MR Egger | 6 | -0.308 | 0.485 | 0.561 |
| | | Weighted median | 6 | -0.370 | 0.272 | 0.174 |
| Stroke | HbF (excluding rs114398597) | Inverse variance weighted (multiplicative random effects) | 5 | -0.410 | 0.083 | 8.17E-07 |
| | | MR Egger | 5 | -0.368 | 0.261 | 0.158 |
| | | Weighted median | 5 | -0.205 | 0.493 | 0.706 |
| ACS | HbF | Inverse variance weighted (multiplicative random effects) | 6 | -0.038 | 0.021 | 0.071 |
| | | MR Egger | 6 | -0.004 | 0.048 | 0.932 |
| | | Weighted median | 6 | -0.043 | 0.022 | 0.044 |
| VOC | HbF | Inverse variance weighted (multiplicative random effects) | 6 | 0.085 | 0.080 | 0.283 |
| | | MR Egger | 6 | 0.309 | 0.174 | 0.150 |
| | | Weighted median | 6 | 0.162 | 0.099 | 0.102 |
| Death | Neutrophil count | Inverse variance weighted (multiplicative random effects) | 247 | -0.594 | 0.478 | 0.214 |
| | | MR Egger | 247 | -0.477 | 0.619 | 0.441 |
| | | Weighted median | 247 | -0.561 | 0.651 | 0.389 |
| Death | WBC count | Inverse variance weighted (multiplicative random effects) | 360 | -0.411 | 0.494 | 0.406 |
| | | MR Egger | 360 | -0.908 | 0.68 | 0.194 |
| | | Weighted median | 360 | -0.669 | 0.744 | 0.369 |
